# A systematic review of the smartphone applications available for corona virus disease 2019 (COVID19) and their assessment using the mobile app rating scale (MARS)

**DOI:** 10.1101/2020.07.02.20144964

**Authors:** Samira Davalbhakta, Shailesh Advani, Shobhit Kumar, Vishwesh Agarwal, Samruddhi Bhoyar, Elizabeth Fedirko, Durga Prasanna Misra, Ashish Goel, Latika Gupta, Vikas Agarwal

**Author notes:** **Corresponding Author:** Latika Gupta, MD, DM, Assistant Professor, Department of Clinical Immunology and Rheumatology, Sanjay Gandhi Postgraduate Institute of Medical Sciences, Lucknow, India. **Funding**- None. **Conflict of interest:** The authors declare that there is no conflict of interest. Code Availability: Not Applicable. **Author Contributions:** All authors were involved with study design, data collection, analysis, manuscript preparation and finalizing the draft.

## Abstract

The global impact of COVID-19 pandemic has increased the need to rapidly develop and improve utilization of mobile applications across the healthcare continuum to address rising barriers of access to care due to social distancing challenges and allow continuity in sharing of health information, assist with COVID-19 activities including contact tracing, and providing useful information as needed. Here we provide an overview of mobile applications being currently utilized for COVID-19 related activities. We performed a systematic review of the literature and mobile platforms to assess mobile applications been currently utilized for COVID-19, and quality assessment of these applications using the Mobile Application Rating Scale (MARS) for overall quality, Engagement, Functionality, Aesthetics, and Information. Finally, we provide an overview of the key salient features that should be included in mobile applications being developed for future use. Our search identified 63 apps that are currently being used for COVID-19. Of these, 25 were selected from the Google play store and Apple App store in India, and 19 each from the UK and US. 18 apps were developed for sharing up to date information on COVID-19, and 8 were used for contact tracing while 9 apps showed features of both. On MARS Scale, overall scores ranged from 2.4 to 4.8 with apps scoring high in areas of functionality and lower in Engagement. Future steps should involve developing and testing of mobile applications using assessment tools like the MARS scale and the study of their impact on health behaviors and outcomes.

## Background

As the Covid-19 pandemic unfolds, it becomes imperative to expand public health activities to estimate the epidemiology for this virus and assess its short term and long term impacts. ^1^ This medical disaster necessitates innovative solutions to address unmet needs as the disease continues to evolve and infect people globally. ^2^ Collection of adequate exposure information, characterizing disease burden and dissemination of information that helps with prevention, containment and contact tracing are vital to supporting public health efforts. In such situations, the use of mobile health applications facilitates self-guided collection of population-level data, delivery of information, addresses knowledge gaps, prevents the spread of misinformation, navigate individuals to accurate and validated health resources. ^3^

Mobile applications provide a unique opportunity in terms of its ability to allow distant connectivity with flexibility of function, design and accessibility. ^4^ Current estimates suggest that the global usage of smartphones stands at 3.5 billion as of June 2020 that provides a large userbase to help coordinate and implement mobile interventions to disseminate and help with pandemic response. ^5,6^ For example, the Singaporean government released a mobile phone app, TraceTogether, that is designed to assist health officials in tracking down exposures after an infected individual is identified. ^7^ Further, Israel passed a legislation to allow the Government to track people with suspected infection through access of their mobile phone data. ^8^ In the U.S. recent legislations have expanded scope of telemedicine to involve use of mobile applications for healthcare and preventive reasons. ^9^ Numerous applications have already been developed globally and are available on the website ((see https://mhealth-hub.org/mhealth-solutions-against-covid-19). In the U.S and UK, the COVID Symptom tracker app developed with the Coronavirus Pandemic Epidemiology Consortium (COPE) is being widely used to track COVID-related exposure and infections. ^10^ However, the downside of this involves access to a large number of applications, some of which can be sources of misinformation or information overload thereby creating barriers to containment and mitigation efforts. Moreover, no universal guidelines exist to ensure quality of these mobile applications to a gold standard.

Our current project aims to provide a systematic review of the literature on mobile applications with multiple function including: information dissemination (ID), ID with symptom checking (SSC), contact tracing alone, SSC, training health care workers, disinfection checklist, and service Apps around COVID-19. Further, we provide an overall rating of the quality of the apps using The Mobile App Rating Scale (MARS) ^11^, a standardized metric, after providing an overview of their functions.

## METHODS

Our systematic review was registered with PROSPERO (CRD 42020180878), consisted of a mixed methods study where we provide an overview of smartphone applications designed for COVID-19 through both a literature search as well as a search of mobile platforms for applications. We describe the app features and assessed their quality in terms of engagement, functionality, aesthetics, and information provided using the MARS instrument.. ^11^

### Search strategy

Mobile applications were searched on the Apple AppStore (AAS), Google Play Store (GPS), National Health Services (NHS) Apps library and Myhealthapps.net. using the keywords-Covid, Corona, Pandemic, Covid-19, SARS-COV2, Symptom, Health, Tracing, Novel coronavirus, 2019-nCoV, and Tracking. We limited our search to applications released or updated for COVID-19 till May 5^th^, 2020 and screened these applications for their title, description of the application on mobile platforms, its functions around COVID-19 and then completed a comprehensive assessment of these mobile applications **(Figure 1a)**. Due to rapidly evolving nature of the COVID-19 pandemic and language restrictions, we limited our search to the United States, United Kingdom and India. Only those smartphone applications with the main subject matter as Covid-19 and in the languages English or Hindi were included in the study. Those applications that were duplicates and pay to download were excluded. Two reviewers each, based across the United Kingdom, United States and India and with access to these applications performed a comprehensive assessment of these mobile applications. Discrepancies regarding selection of apps were resolved by a third reviewer in order to reach a consensus.. (Supplementary Table 1).

**Figure 1a:**
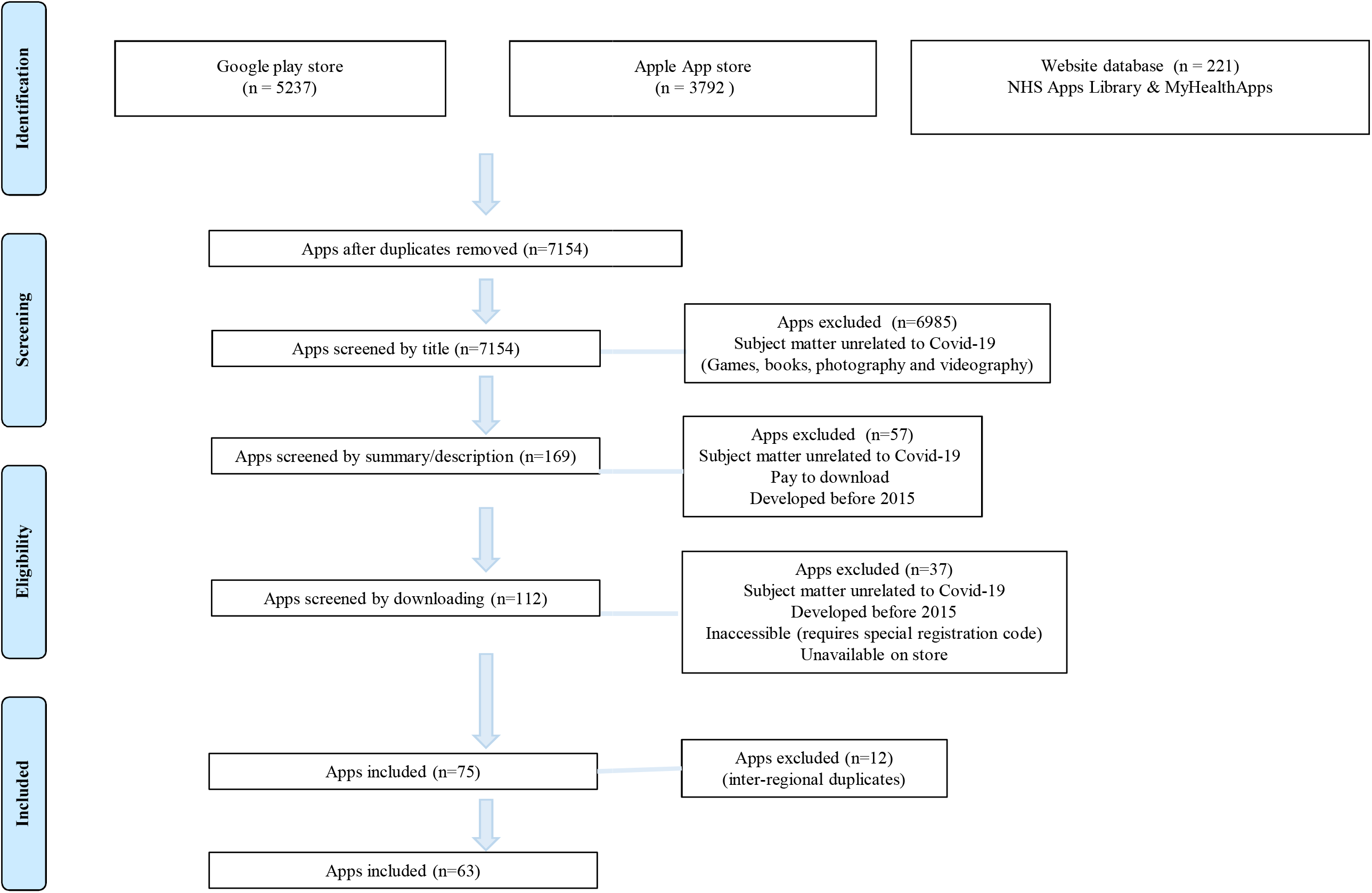
PRISMA-Mobile app search

**Figure 1b:**
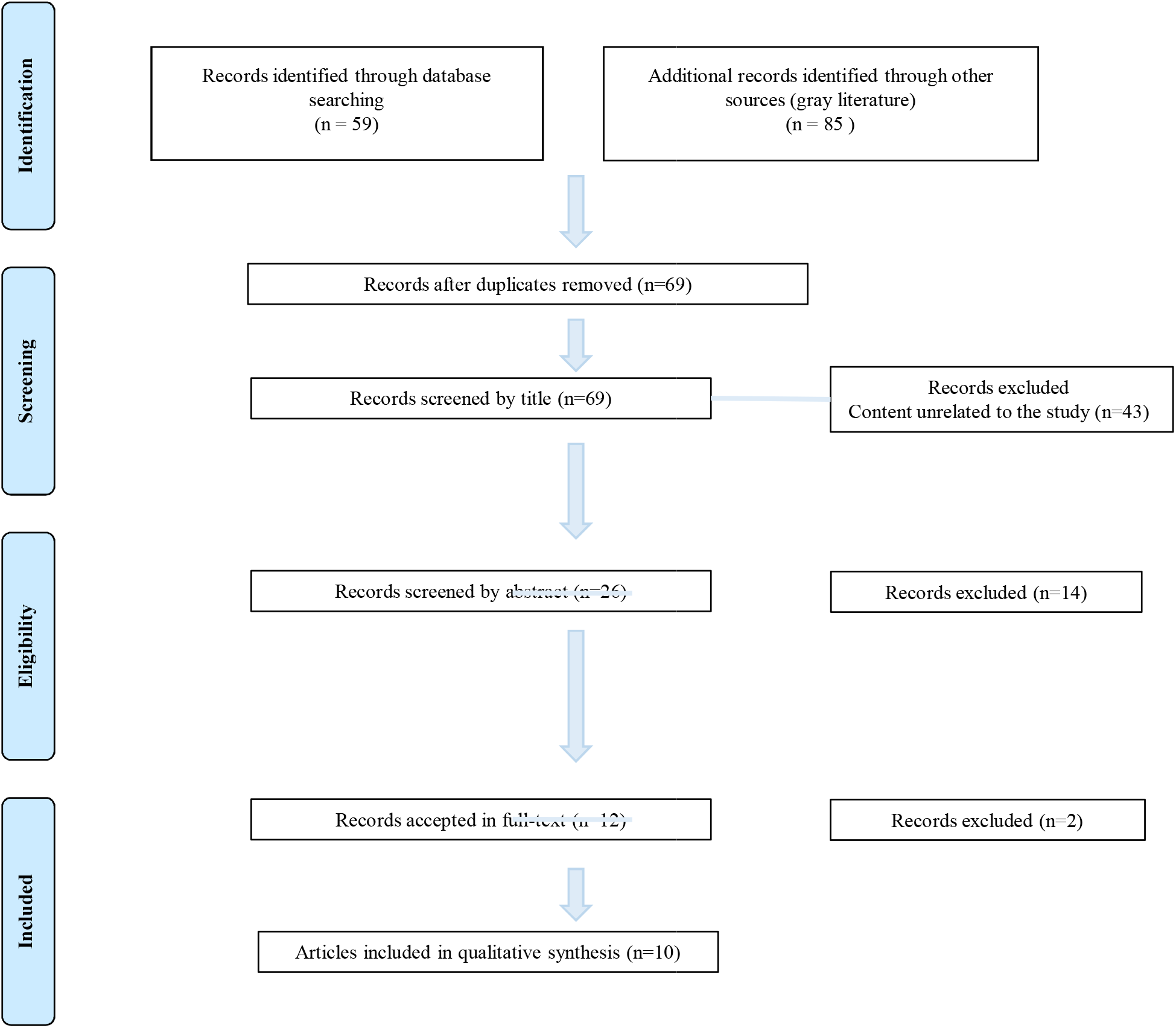
PRISMA-Literature search

Subsequently, data was extracted from each app (Supplementary **Table 2**) and each app was assessed using the MARS rating scale. ^11^ The MARS evaluation tool is divided into three broad sections-App overall quality, App subjective quality, and App specific quality. It consists of twenty-three questions that are designed to help analyse engagement, functionality, aesthetics, information, and subjective quality of the mobile applications. In addition, there are six final app specific questions that can be tailored to represent the target health behaviour/ function of the application/study (**Figure 2**). The mean scores for each of the four subscales (Engagement, Functionality, Aesthetics, and Information) was calculated, and the mean scores of those subsections, used to rate the total quality score of the app ranged from zero to five. Hence the overall score on these subscales would range from 0 to 5 and we would develop an average score by dividing the total sum score by 4 i.e. 4 domains. The App subjective quality section and App specific section was calculated similarly. These, however, were regarded separate from the app quality score. All reviewers underwent training in the use of MARS using a 37-minute video available on YouTube ^12^ which was followed by an independent review exercise of a common app and discussion of scores to assess consensus. Disagreements were discussed with a third reviewer, and ambiguous MARS items were clarified to ensure full comprehension of the scale. ^11^

**Figure 2:**
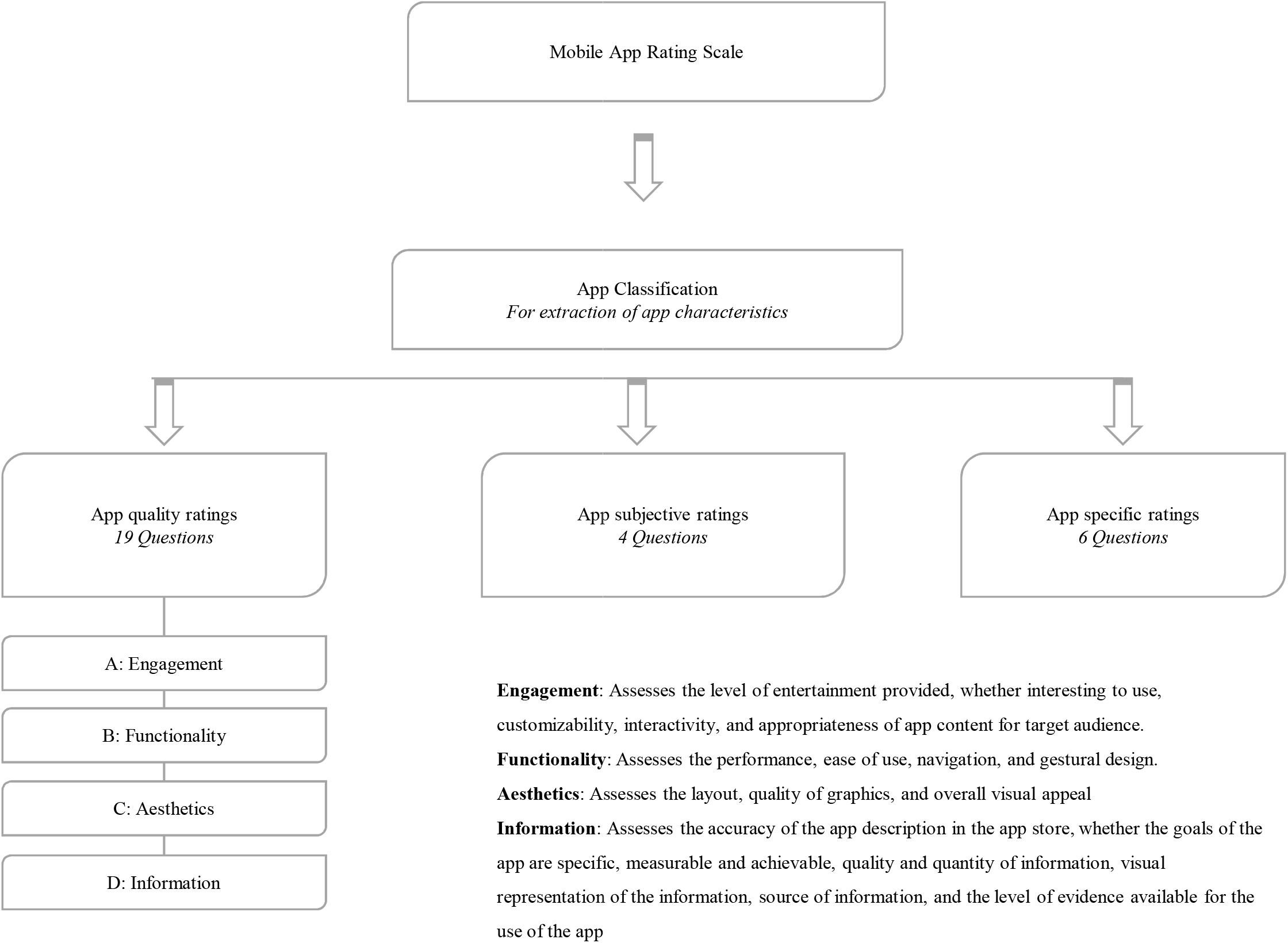
MARS structure

**Figure 3:**
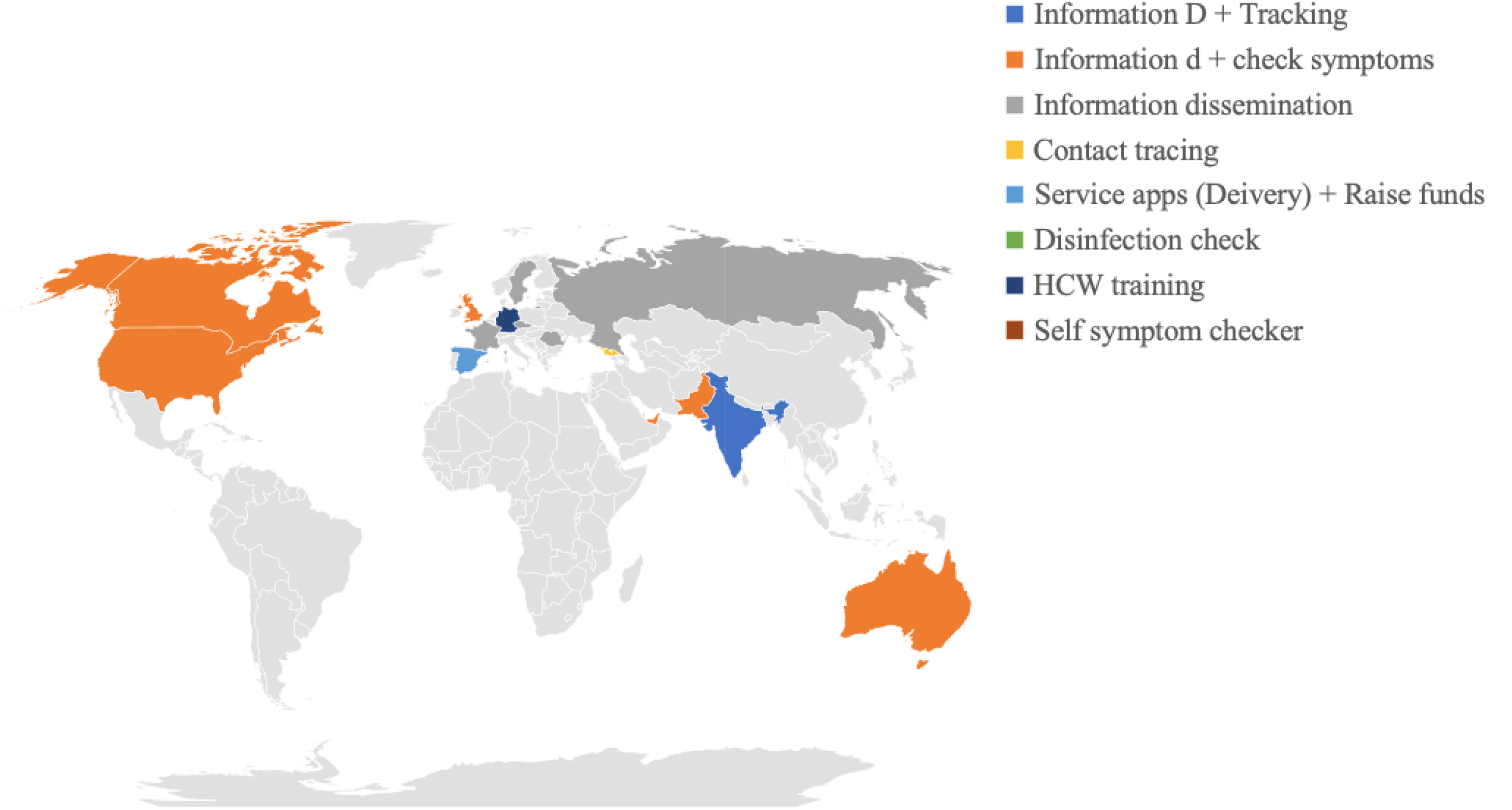
Map of country of origin

### Statistical analysis

Unpaired students t-test was performed to analyse the difference in the means obtained in each section by Asian, European, and North American apps with a p value of <0.05 considered as significant. Pearson’s correlations were used to analyse correlations between app downloads and MARS ratings. Microsoft excel was used for data segregation and analysis.

#### Literature search

A systematic search was conducted on PubMed (Medline), Cochrane central register for controlled trials, Google Scholar and Turning Research Into Practice medical database from 9-10^th^ of May 2020 to identify peer-reviewed publications and grey literature on the utility of smartphone applications in the Covid-19 pandemic and help aid our ongoing assessment of mobile platforms. ^13^

The screening process was conducted by two review team members using the keywords: “Covid”, “Corona”, “Pandemic”, “Covid-19”, “SARS-COV2”, “Novel coronavirus”, “2019-nCoV”, and “smartphone”. The articles were screened for relevance by title, abstract and eventually full-text review was performed for included abstracts (**Figure 1b**). Articles in languages other than English were excluded. Those studies in which the subject matter was unrelated to the use of smartphone apps in Covid-19 were excluded.

## RESULTS

Our search identified 63 apps that were reviewed and performed a quality assessment on using the MARS scale. Of these, 21 apps originated from Asia, 17 from Europe, 22 from North America, 2 from Australia, and 1 from Russia (**Supplementary Figure 1)**. Apps were classified based on their key functions-ID (n=18), ID with tracking of individuals (n=9), or ID with SSC (n=15), contact tracing alone (n=8), SSC (n=7), training of healthcare workers (n=3), disinfection checklist (n=1), and service apps [Delivery or fund-raising] (n=2).

### An overview of Characteristics of apps

Of all the apps, 27 (39%) were developed through federal efforts, 14 (21%) by commercial units, 5 (7%) by a University, 1 (1%) by an NGO and 16 (24%) by unknown organisations/ entities (**Supplementary Table 2**). Data extraction for the number of downloads of the app was only possible on the android apps as that information is not available on the Apple app store (AAS). The average star rating of apps on both the stores was 4.2 (±0.07)

The number of app downloads ranged from 10+ to 50,000,000+. The most downloaded and reviewed app on the Google play store (GPS) was *Aarogya Setu* developed by the Government of India (50,000,000+ downloads and 343,842 reviews), with a government mandate for download by all Indians especially those who travel or are diagnosed with COVID-19, with a GPS star rating of 4.6 out of 5 (Supplementary Table 2). In addition to providing self-screening technology, it displays alerts of COVID-19 patients diagnosed within a 500 meter, 1, 2, 5, and 10-kilometre radius of the user which is a useful tool to understand the disease load in the proximity of an individual. Average star rating on play store was 4.2 (±0.5).

### Mobile Applications Overview

#### App quality assessments and ratings using MARS

The mean overall MARS score obtained was 3.7 of 5 (SD=±0.58). Of 63 applications, 23 apps had a score above and equal to 4, most of which were from the information with symptom checker (8), information with tracking (6), and purely information dissemination (5) categories. Eleven apps had a score of less than 3 (**Table1)**. The highest rated app was *Covid-19 UAE* (4.8) closely followed by *First Responder Covid-19* (4.8) and the lowest score was for *Corona 360* (2.4).

All the apps were initially assessed using MARS for Engagement, Functionality, Aesthetics and Information mean scores for each subscale (out of 5) calculated (details in Figure 2). Overall, apps scored highest in the functionality (4.5, SD=0.6) domain, followed by aesthetics (3.8, SD=0.8), information (3.5, SD=0.8) and engagement (3.1, SD= ±0.8).

*Covid-19 UAE* scored a 5 in the Engagement, Functionality, and Aesthetics and 4.2 in the information sub-scale. In comparison, *Corona360* scored 1.6 in engagement, 3.3 in functionality, 3.3 in aesthetics and 2 in information. *Corona360* is an app developed by a private company, which aimed to create a map based visual database of individuals infected with COVID-19 by allowing anonymous sharing of the users’ status of infection along with their approximate location. This app lacked methods of customisation or user engagement. The singular graphic used in the app for a map with locations of infected individuals made it difficult to use and slow to load, which might have contributed to low engagement.

Apps from Asia scored higher in functionality (mean difference 0.54 (± 0.13) 95% CI= 0.3 to 0.8, p= 0.0001), while the UK (8 of 17 from Europe) and North American apps together scored higher in the information subscale (mean difference 0.6 (± 0.2) 95% CI= -1.0 to -0.1, p= 0.01). Aesthetics, engagement and total scores did not differ between the western and Asian Apps. Thus, the apps based in the Indian subcontinent were sufficiently interactive and functioned smoothly, but lacked in either quality, or quantity of information provided. In comparison, those apps developed in North America of the United Kingdom had credible, necessary information, but lacked in creative methods of presentation of this information which could be understood by the masses. A detailed analysis into each sub-scale was performed using the components of the MARS scale^11^ as previously described:

##### 1) Engagement

Two apps scored a 5 in this section-*First Responder* and *Covid-19 UAE* while Pakistan’s National Action Plan scored the lowest, with a score of 1.4. *First Responder* was developed by Stanford University and designed especially for healthcare workers to screen their symptoms and, if needed, schedule a testing appointment at Stanford Health Care. The app has a well-chosen colour scheme. Key features of this app included the use of pictorial representations of healthcare workers admixed with color-coded sections that make it easy for the users to navigate through different sections. It increased its level of interaction with the user by the self-screening feature of the app, wherein the user can assess their health themselves and be given further guidance as to whether they should self-quarantine or go to a health care provider. Enough customisation was present in the form of an edit function in self-screening history and the notification settings.

The *Covid-19 UAE* developed by the Ministry of Health UAE’s official mobile app on Coronavirus management provides users with real-time Coronavirus case information and keeps the app users engaged by sharing news up to date with regional news and events. Its content presentation includes eye catching graphics such as use of cartoon graphics representing viruses on top of each section. The app gives a color-coded description of current status of patients: including newly diagnosed cases, deaths and those that recovered. Moreover, it allows customisation to three different languages, including English, and a self-assessment section for Covid-19.

Pakistan’s National Action Plan for Covid-19 was created by a private company and aimed to provide information on the steps the country will take to tackle Covid-19. This app scored low in this section as it lacked any form of graphics altogether. The information was present as a PDF with the only interaction being the ability to change the pages and did not engage its users overall.

##### 2) Functionality

Twenty-eight apps scored a 5 in the functionality sub-scale. Of these, *Jaano* and *COVA Punjab* worked with appropriate speed and switching between sections of the app was smooth. The menu labels and icons were interactive and clear instructions and navigations made apps like *Babylon-Healthcare service* and *NHS24:Covid-19* easy to use and easily comprehendible.

As for those apps which scored the lowest (<2.5), the main operation of the app did not work appropriately. For example, if the purpose of the app was to provide a map of infected individuals in the locality, the map itself would not open.

##### 3) Aesthetics

*T COVID’19* and *Odisha Covid Dashboard* both available on the GPS and AAS, scored a 5 in this section. *T COVID’19* developed by Government of Telangana, India from provides updates regarding the cases and deaths due to Covid-19, information about the nearest health centres, essential services, and Government announcements, and allows self-symptom checks and tracking of infected individuals. The salient feature of this application involves use of interactive icons used to depict different sub-sections of the app; for example, the icon for ‘telemedicine’ included a doctor with a chat box next to him. *Odisha COVID Dashboard* utilises colour codes to depict the main sections of the app, which increases its visual appeal. Each graph is explained carefully and clearly with the colours to depict the case burden i.e. red, orange, green and grey depicting confirmed, active, recovered and deceased cases respectively. Further the map provides a heatmap of the State to represent Covid-19 infection burden across state districts. The graphics used were overall of a high quality. Further, the app posts important announcements including health advisories in forms of health posters.

The apps scoring the lowest in this section (2.3) intended to track the user’s location and alert of a possibility of exposure. Poor aesthetics, including low quality graphics, and small buttons and icons made navigation difficult, and hampered the ability to perform its main function.

##### 4) Information

The highest rated app in the information sub-scale was *1-Check COVID* (4.8) followed closely by *NHS24: Covid-19* (4.75). *1-Check COVID* was developed by health experts from the University of Nebraska Medical Center. It has an accurate and precise description on the app store, which clearly depicts the goals, i.e. providing a real-time situational awareness of Covid-19 and public health risk assessment survey. The app asked for details about the users’ symptoms, travel, medical conditions, and exposure with clarity, and provided credible health information links (e.g. Centre of Disease Control) to help the user learn more about Covid-19 features. Further, the NHS:24 app provides appropriate advice based on a simple symptom assessment. It may advise its users to call an NHS number (111) or advise self-isolation for a period, based on NHS guidelines. This information is easy to comprehend and written in simple and easy language. Besides, numerous sections covered a range of topics, and many of these contained further links to other reliable websites (e.g. NHS Inform). The Information was presented under a clear heading, categorised into sections e.g. General Advice, Testing and Physical Distancing.

### App subjective quality items and App specific items (Figure 2)

The median app subjective quality score was 3.25 (±1.01,). Top and lowest scoring apps in the specific items are summarized in **Table 1**.

### Correlation between MARS app quality score, and the number of downloads/ app star rating

The MARS scores and number of downloads did not correlate [*r*^(2)^ = 0.082, *p* = 0.65]. No correlation was found between the MARS score and app star ratings either [*r*(^2)^ = 0.064, *p* = 0.64]. However, a significant correlation was found between app star ratings and MARS scores for those apps used in North America [*r*(2) = 0.6, *p* = 0.006].

### Literature Search

We identified 144 articles based on our search strategy (Figure 1B). After reviewing our inclusion and exclusion criteria, 134 research studies were excluded, majority of which were due to their content being unrelated to our study. For example, those articles which focused on specialty practice during COVID-19 and telemedicine, smartphone enabled technology that did not involve an application, or studies that utilized apps to assess for mental health outcomes among COVID-19 patients.

A total of ten studies were found on this subject and have been detailed in **table 2**. Majority of ongoing research focused towards development of smartphone app technology for contact tracing with the preservation of user data, apps for information dissemination to healthcare workers in a region (n=1), and smartphone app enabled technology for diagnostics. The more descriptive studies reviewed the evidence of the utility of smartphone apps for COVID-19 based on countries’ experiences (**Table 2**). Only one application included in our study, The *COVID-19 Symptom Tracker* mobile application, conducted an observational study to provide scientific evidence of the app’s utility. Through the application, they were able to acquire important information regarding the population dynamics of the disease amongst the users. Comparisons between symptoms of users and test results allowed generation of hypotheses for further research. Important results in their study demonstrated that the app could predict in advance the COVID-19 incidence of a region. ^14^

## DISCUSSION

To our knowledge, our study is one of the first to perform a comprehensive assessment of mobile applications being used during the COVID-19 pandemic. Smartphone applications have an immense potential to control the spread of misinformation associated with the COVID-pandemic, raise awareness, allow contact tracing and finally, help improve provision of the preventive and clinical care of patients. We identified 63 mobile applications from different regions which scored across the MARS continuum and differed in terms of their functionality, aesthetics, information sharing and overall purpose. The app downloads did not correlate with quality, suggesting the need to revisit the app usage strategies among individuals and advisories by federal or health authorities.

Our results identified 63 apps that are currently being utilised across different arenas of the pandemic. Overall, the apps analysed by us demonstrated above average quality. Most (53) of the apps acquired 5/5 in functionality of which only one fifth (12) apps scored similarly in Engagement. This shows that most apps focus on their functioning without considering features that make the app equally engaging and important to an increased userbase. Moreover, the large number of apps delegated by various countries and organisations could potentially cause confusion amongst users regarding the utility and preference. The fact that the app quality did not correlate with downloads supports this possibility. Moreover, extending the usage of quality apps designed in one region to others may be a cost-efficient and relevant administrative exercise in the face of a global economic collapse. We identified *Covid-19 UAE, NHS24:Covid-19, Odisha Covid Dashboard*, and *1-Check COVID* as exemplary apps in the engagement, functionality, aesthetics, and information domains respectively.

Interestingly, the apps developed in Asia were user-friendly and designed to ensure that most of the users understood the application and used it effectively. With over 500,000,000 smartphone users in this country^15^ and considerable variations in socioeconomic status including differences in education attainment these characteristics are extremely necessary to be considered in developing a user-friendly app. On the other hand, the apps accessed from Europe/ North America remained focused on information. Though high quality and from credible sources, they often lacked creative or interactive methods to portray this information. This may potentially decrease the assimilation of the knowledge amongst the users, defeating the purpose.

Besides, apps developed in the west tended to focus on information dissemination, with very few contact tracing or symptom checking apps. On the contrary, most apps developed in India have been created with the main purpose of contact tracing, or symptom checking. Emerging events of data leaks and technological failure have raised heightened concerns over data-safety laws which range from very stringent (e.g. Germany) to blurry, and almost non-existent in some countries. In a time with abundant anxiety and chaos alongside over uncertainty about the future, this could be a recipe for disaster.

‘*Aarogya Setu’* a contact tracing app developed by the Government of India ^16^ has 50,000,000+ downloads. It’s ability to function adequately and provide credible information make it a suitable app to reach out to the large population of India. However, the requirement of constant blue-tooth access and location tracking, has been perceived as a violation of privacy, more so as it lacks the mandatory layers of system privacy expected by an App promoted at such a scale. ^17,18^ The use of centralized digital contact tracing is debatable. Governments like France, UK, and Italy favour centralization wherein their public health authorities receive information immediately, about contacts of infection. Others including companies like Google and Apple prefer a decentralized approach. Here, the contacts are notified of proximity to an infected individual and can choose to share this information with health authorities. Recently a mandate to download the Aarogya Setu app for screening in public spaces received harsh criticism from proponents of ethics and cyber-laws. ^19^ Although the use of smartphone applications comes with numerous concerns regarding data privacy, studies have shown that it may be possible to use apps to fight Covid-19 while preserving an individual’s right to privacy.^20,21^

Repurposing can deliver useful and quality apps in a timely fashion, with the added advantage of accessing active users who have already downloaded the app. Apps such as *Babylon-Healthcare service, CDC, WebMD, and MyGov* have been repurposed for COVID-19. Updating these to perform functions like contact tracing could save time and effort. Moreover, merely 24 of the 63 apps assessed by us had overlapping functions (Information dissemination with self-symptom checker/ contact tracing). The presence of most necessary functions in one place is essential to facilitate a wide usage of the app.

Research into smartphone applications for Covid-19 is sparse. Despite the plethora of Apps in the market, our literature search identified only ten studies pertaining to this subject. Most of them demonstrated that the development of a privacy preserving contact tracing technology enabled by a smartphone app is possible. Two studies focused on the utility of smartphone apps in diagnostics for Covid-19 patients, advocating its use due to its ease and self-diagnostic nature. Patients in need of heart rate monitoring, or those requiring Covid-19 testing could possibly use this novel technology themselves and report it to a healthcare provider. Future Randomised Controlled Trials (RCTs) using mobile applications for Covid-19 and its impact on public awareness, disease spread, and healthcare delivery are warranted. Health apps advocated for the protection of users during a global pandemic such as this must be backed by scientific evidence, without which a strong hold on the quality and utility of the app is bound to be missing.

The psychometric properties of the MARS scale are proven to be reliable and valid, thus the use of this tool provides great strength to our study. ^11^ With a large range of keywords and an additional literature search, our study has a very wide reach by using a mixed methods approach. Further, our study provides a comprehensive assessment of all mobile applications across three regions which are currently experiencing rapidly rising burden of COVID-19 related cases and deaths and hence, identifying the key scientific features of mobile applications that should be used in design and engagement remains very crucial to sustained public health efforts aimed at mitigating this disaster. However, apps in languages other than English and Hindi could not be assessed, which limits the generalizability of our results to other regions. Despite a high interrater reliability ^11^ the reviewer’s subjectivity might have influenced the ratings awarded and caution must be exercised while interpreting the results portrayed in this review. In addition, there is a likelihood that the features of the apps portrayed by us are different from the updated versions of the app and might have been addressed in apps developed after this review. This possibility is inevitable considering the rapidity with which apps are developed and reformed.

## CONCLUSION

In the recent Covid-times, the market has seen the appearance of numerous mobile-applications for tracking and managing the pandemic. The app-market remains disorganized and unregulated in several countries. The present review provides an overview of mobile applications available in United Kingdom, USA and India; summarize their strengths and limitations through a qualitative assessment; and delineates key functions and features needed for future applications. Rapid population-based longitudinal studies and randomized trials would characterize the use and efficacy of mobile apps on health knowledge, behaviours and use to limit the spread of COVID-19 and help reduce its burden on the public health and clinical systems.

## Data Availability

All data is available as tables and figures submitted along with the manuscript.

## Legends to figures and tables

Table 1: App quality ratings

Table 2: Literature search results.

## Supplementary files

Supplementary Table 1: Discrepancies resolved while eliminating apps by name and abstract review

Supplementary Table 2: Characteristics of apps assessed using the MARS

## Acknowledgements

Michael Beadle, Arya Ghatge-Participated in the mobile application screening and rating process.

